# An economic evaluation of Kooth, a web-based mental health platform for children and young people with emerging mental health needs

**DOI:** 10.1101/2022.08.12.22276381

**Authors:** Laura Coote, Laura Kelly, Louisa Salhi, Santiago de Ossorno Garcia, Aaron Sefi, Hayden Holmes

## Abstract

**Background:** The Kooth platform is a mental health web-based platform commissioned by the NHS, local authorities, charities and businesses. Through the platform children and young people (CYP) have access to an online community of peers and a team of experienced counsellors.

**Objective:** To evaluate the potential benefits of the Kooth mental health platform in the UK, including the costs and savings to public services and impact on both immediate and intermediate outcomes for CYP.

**Methods:** A cost calculator was built to estimate the cost to the UK government as a result of implementing the Kooth platform. A decision tree structure was used to track the progress of CYP with emerging mental health needs (EMHN), comparing those with access to the Kooth platform and those without access to a mental health platform.

**Findings:** The base case results followed a cohort of 2,160 CYP. The results of the cost calculator estimated that engagement with the Kooth platform is associated with a cost saving of £303,234 or £199 per engaged user, to the NHS and UK crime sector.

**Conclusions:** The uptake of mental health platforms, such as Kooth, could result in cost savings to the UK public sectors and should be given consideration as a beneficial addition to existing mental health services.

## Background

One in six UK school-aged children has a mental health problem. A rise from the one in ten reported in 2004 and the one in nine in 2017 (1). Mental health problems in adolescence are reported to result in a greater risk of physical and mental health problems in adulthood (2). Mental health problems in young people are associated with risky behaviours, such as smoking, drug and alcohol abuse (3).

Child and Adolescent Mental Health Services (CAMHS) are a part of the UK National Health Service (NHS), they assess and treat young people with emotional, behavioural or mental health difficulties (4). CAMHS comprises of local services around the UK, with teams made up of nurses, therapists, psychologists, child and adolescent psychiatrists, support workers and social workers, as well as other professionals. CAMHS has come under increasing pressure, with a rise in referrals alongside a growing number of young people presenting to emergency departments (5). The Education Policy Institute revealed that 26% of referrals to specialist CAMHS were rejected in 2018/19 (5). Not meeting the eligibility criteria was the most frequent cause for rejection (6).

Kooth.com is a mental health web-based platform commissioned by the NHS, local authorities, charities and businesses. The platform is intended to give children and young people (CYP) easy access to an online community of peers and a team of experienced counsellors.

## Objective

To evaluate the potential benefits of the Kooth mental health platform in the UK, including the costs and savings to public services and impact on a range of immediate and intermediate outcomes for CYP.

## Methods

Modelling was undertaken to estimate the costs to public services in a population when Kooth is available, compared to when Kooth is not available. To measure the benefits of Kooth, we calculated outcomes such as the number of hospitalisations due to suicidal ideation and self-harm, crimes committed, smoking status, binge drinking, the number of GP appointments and antidepressant prescribing. The perspective was public services in the UK, specifically the healthcare and crime sector. All costs are reported in UK pounds at 2019/20 price levels. The time horizon was one year and there was no discounting.

### Population

The study population of interest were children and young people (CYP) with emerging mental health needs (EMHN) in the UK. The Young Person’s CORE (YP-CORE) questionnaire was used to determine the proportion of CYP with an EMHN. The YP-CORE is a measure of psychological distress and is widely used in mental health and school counselling services (7). A score of 15 (moderate level) or greater on the YP-CORE at Kooth registration was used as a proxy for the EMHN population. A cohort of 2,160 people was used as the model population. This was based on a current contract between Kooth and a local authority in England.

### Model structure

The model structure was separated into two parts. Firstly, a decision tree was used to determine the proportion of the registered CYP population engaging with the Kooth platform, as seen in Figure 1. Engagement was defined as young people who create a Kooth account and then complete an activity on the platform within one month. The proportion of those engaging were taken from data provided by Kooth. From October 2019 to June 2021 the engagement rate was 76.5%.

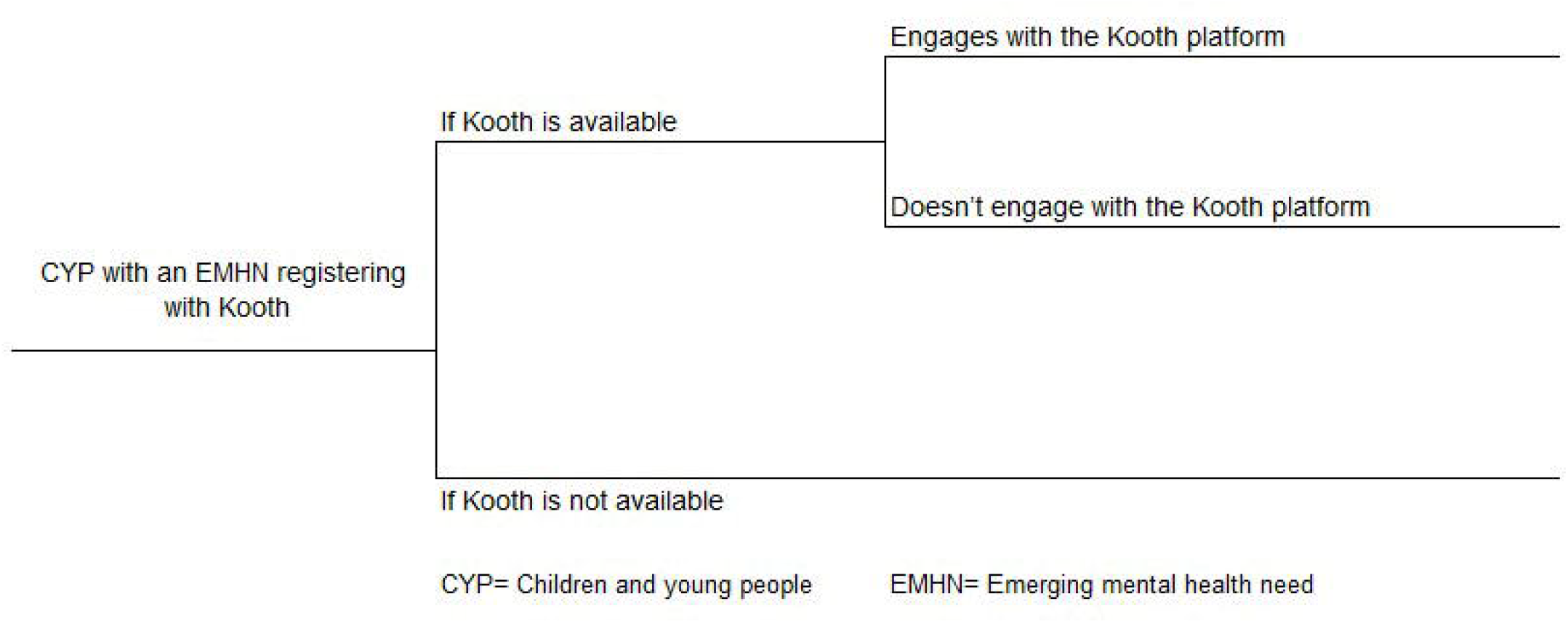

Changes in immediate outcomes of young people, as a result of engaging or not engaging with the Kooth platform, were taken from a study of Kooth published by the London School of Economics (LSE) in 2021 (8). The study used online baseline and follow up surveys to assess the mental health and wellbeing of Kooth registrants. Baseline scores were used as the outcomes for young people not engaging with Kooth. One-month follow-up outcomes were used as outcomes for young people engaging with the service. These outcomes are termed immediate outcomes in the model and include:

- Suicidal ideation (Suicidal Ideation Attributes Scale [SIDAS]) (9).
- Self-harm (a2-item questionnaire) (10).
- Perceived impact of difficulties (Strengths and Difficulties Questionnaire [SDQ]) (11).

The model assumed that the service could only impact the immediate outcomes of young people who had previously presented with these outcomes to a Kooth practitioner. This could be either via a 1-1 therapeutic chat, or interaction with the moderated Kooth community. These Kooth collected ‘presenting issues’ were mapped to the LSE studied immediate outcomes via a categorisation exercise workshop between Kooth and three of Kooth’s internal clinical experts. The presenting issues of Kooth users were as follows, 25.3% with suicidal ideation, 26.2% self-harm and 71.2% with perceived impact of difficulties (SDQ).

The second part of the model structure linked a change in immediate outcomes to a set of intermediate outcomes, see Figure 2. Intermediate outcomes are societal events associated with CYP mental health in which a cost per event can be calculated. These outcomes included the number of hospitalisations due to suicidal ideation and self-harm, crimes committed, smoking status, binge drinking, the number of GP appointments and antidepressant prescribing. The difference in the number of intermediate outcome events, with and without the availability of Kooth to CYP in the UK, is calculated as a model outcome. Immediate outcomes were linked to intermediate outcomes using odds ratios (OR) and relative risks (RR), found in the literature. Table 1 outlines the OR and RR used, as well as the baseline prevalence of intermediate outcomes found in CYP.

**Table 1.** model inputs

| **Difference in scores between baseline and follow up** |  |
| --- | --- |
| SDQ | -2.2%(8) |
| SIDAS | -8.8%(8) |
| Self-harm | -19.2%(8) |
| **Population** |  |
| Kooth engagement rate | 76.5% |
| Proportion of Kooth users with an EMHN | 92% |
| **Presenting issues of Kooth users** |  |
| Suicidal Ideation | 25.3% |
| Self-harm | 26.2% |
| Perceived impact of difficulties (SDQ) | 71.2% |
| **OR/RR linking immediate and intermediate outcomes** |  |
| Suicidal ideation (SIDAS) to Hospitalisation OR | 3.75 (15) |
| Self-harm to Crime OR | 3.5 (16) |
| Self-harm to Smoking RR | 2.21 (17) |
| Self-harm to binge drinking RR | 2.13 (17) |
| SDQ to smoking OR | 1.14 (18) |
| SDQ to binge drinking OR | 2.18 (19) |
| **Baseline prevalence of intermediate outcomes** |  |
| Suicide attempt (SA) resulting in hospitalisation | 8.46%(20, 21) |
| Arrests | 2.11% (22) |
| Regular smokers 11-15 years | 2.0% (23) |
| Regular smokers 16-24 years male | 23.0% (24) |
| Regular smokers 16-24 years female | 19.0% (24) |
| Binge drinkers | 28.0% (25) |
| Percentage of EMHN going to visit a GP | 75.0%* |
| Percentage of EMHN who visit their GP that will have monthly GP follow ups, an antidepressant prescription and CAMHS referral | 20.0%* |
| Percentage of CYP who are referred to CAMHS that will be rejected and go back to the GP | 25.0%* |
| **Costs** |  |
| Hospitalised (SA population) | £877† (26) |
| Arrests | £1,026† (27) |
| Regular smokers 12-24 years | £257‡ |
| Binge drinkers | £701‡ |
| Example cost of a Kooth contract | £140,000 |
| Cost per GP appointment | £33.00 (12) |
| Cost of 1 year of Fluoxetine | £751.56 (13) |

CYP, Children and young people; EMHN, Emerging mental health needs; OR, Odds ratio; GP, General practitioner; RR, Relative risk; SA, Suicide attempt; SDQ, Strengths and Difficulties Questionnaire; SIDAS, Suicidal Ideation Attributes Scale.
\*Assumption validated from clinical experts.
†Inflated to 2019/20 prices
‡ For full calculation details see Supplementary data.

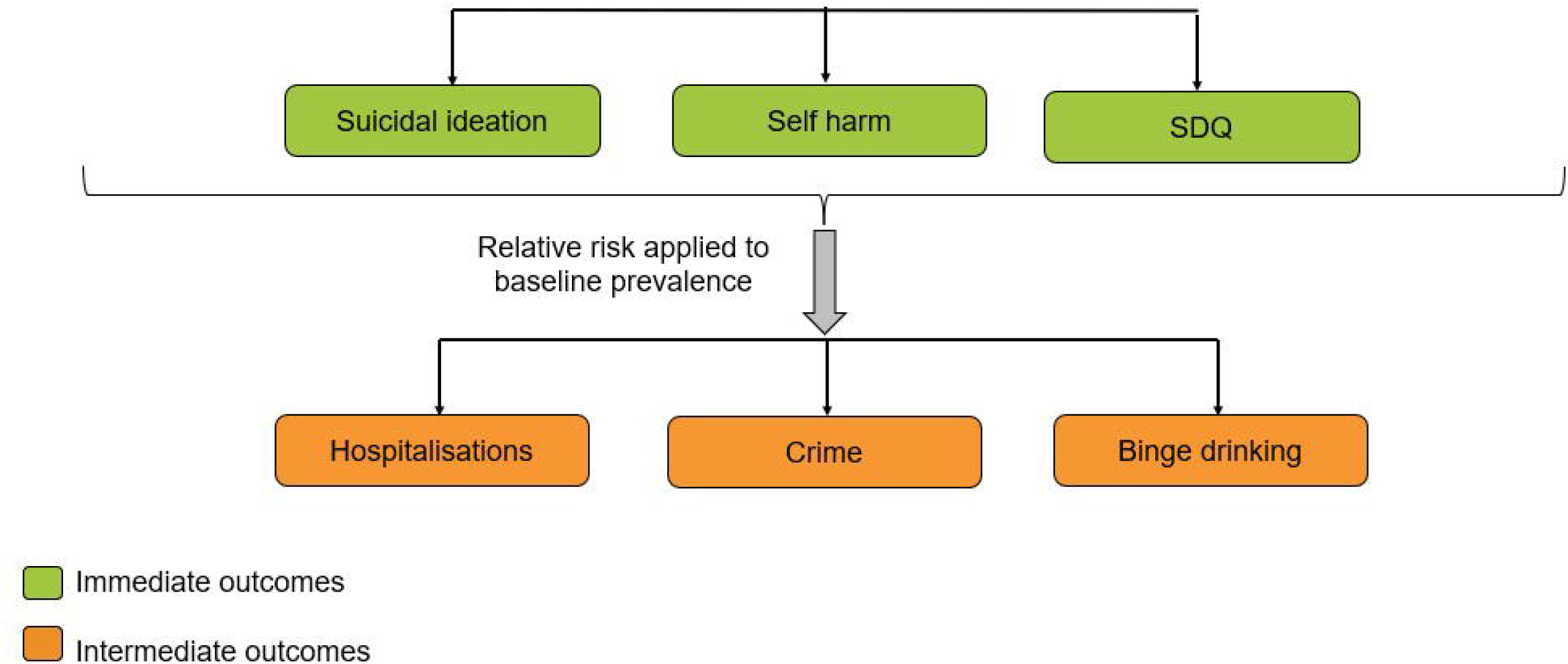

It was assumed that 75% of CYP with EMHN visit their GP, and 20% of those have monthly GP appointments, are prescribed antidepressants and referred to CAMHS. Furthermore it was assumed that 25% of CYP referred to CAMHS were rejected and return to their GP for help. The above assumptions were validated by clinical experts.

### Costs

Costs used in the model are detailed in Table 1. Costs were obtained from a targeted literature search. All costs are reported in UK pounds at 2019/20 price levels. We followed published guidance on exchange rate adjustment and accounting for inflation (12).

The cost of medication used in the model was the cost of 30 days on Fluoxetine of 10mg daily, scaled to 1 year (13). Fluoxetine is licensed in the UK for children and adolescents aged 8 years and over for treatment of moderate to severe depression. The electronic medicines compendium (EMC) reports the starting dose is 10mg either as 2.5ml fluoxetine liquid daily or as one 20mg capsule on alternate days, increased to 20mg (usually capsules) after 1-2 weeks if necessary (14). Where most people take fluoxetine for at least six to 12 months after they start to feel better (4).

The cost of providing Kooth services varies depending on area and number of users. For the base case a value of £140,000 was used. This value was taken from a previous Kooth contract for a local area of 2,160 young people in England.

### Sensitivity analysis

Deterministic sensitivity analysis (DSA) was undertaken to evaluate the impact of input assumptions on the cost difference between young people having access to Kooth or not. In DSA, input values varied between the lower and upper bound of its 95% confidence intervals. When a confidence interval was not reported, ±20% of the baseline value was used. The contract cost upper and lower values were taken from a contract costs provided by Kooth. All analyses were conducted using Microsoft Excel. We followed published guidance on the reporting of economic evaluations(28).

## Findings

The base case results follow a cohort of 2,160 CYP with EMHN. The results of the cost calculator estimated that engagement with the Kooth platform is associated with cost savings that outweigh the cost of the Kooth contract. A cost difference of -£303,234 or -£199.48 per engaged user, to the NHS and UK crime sector is shown in Table 2. These costs are broken down further in Table 2, showing that the biggest cost saving was due to the reduction in antidepressant prescribing, which resulted in an overall cost saving of £223,966.

**Table 2.** Results of economic model

| Overview | Total | Per engaged EMHN CYP |
| --- | --- | --- |
| Cost of Kooth | £140,000 | £92.10 |
| Cost saving | £443,234 | £291.58 |
| Cost difference | -£303,234 | -£199.48 |
| **Intermediate outcome** | **Reduction in incidence** | **Cost saving** |
| Hospitalisations averted due to suicidal ideation | 6 | £5,262 |
| Hospitalisations averted due to self-harm | 13 | £11,401 |
| Crimes averted | 3 | £3,078 |
| Smokers averted | 39 | £10,256 |
| Binge drinkers averted | 28 | £19,618 |
| GP appointments averted | 5,141 | £169,653 |
| Antidepressant prescriptions averted | 298 | £223,966 |

CYP, Children and young people; EMHN, Emerging mental health needs.

### Sensitivity analysis

The robustness of the model was tested using DSA, the findings are shown in Figure 3. The DSA results show that the parameter most affecting the cost difference per person is the proportion of CYP who engage with a GP. It is assumed that those who engage with Kooth do not require GP appointments for their mental health. For all input value variations, Kooth remained cost saving.

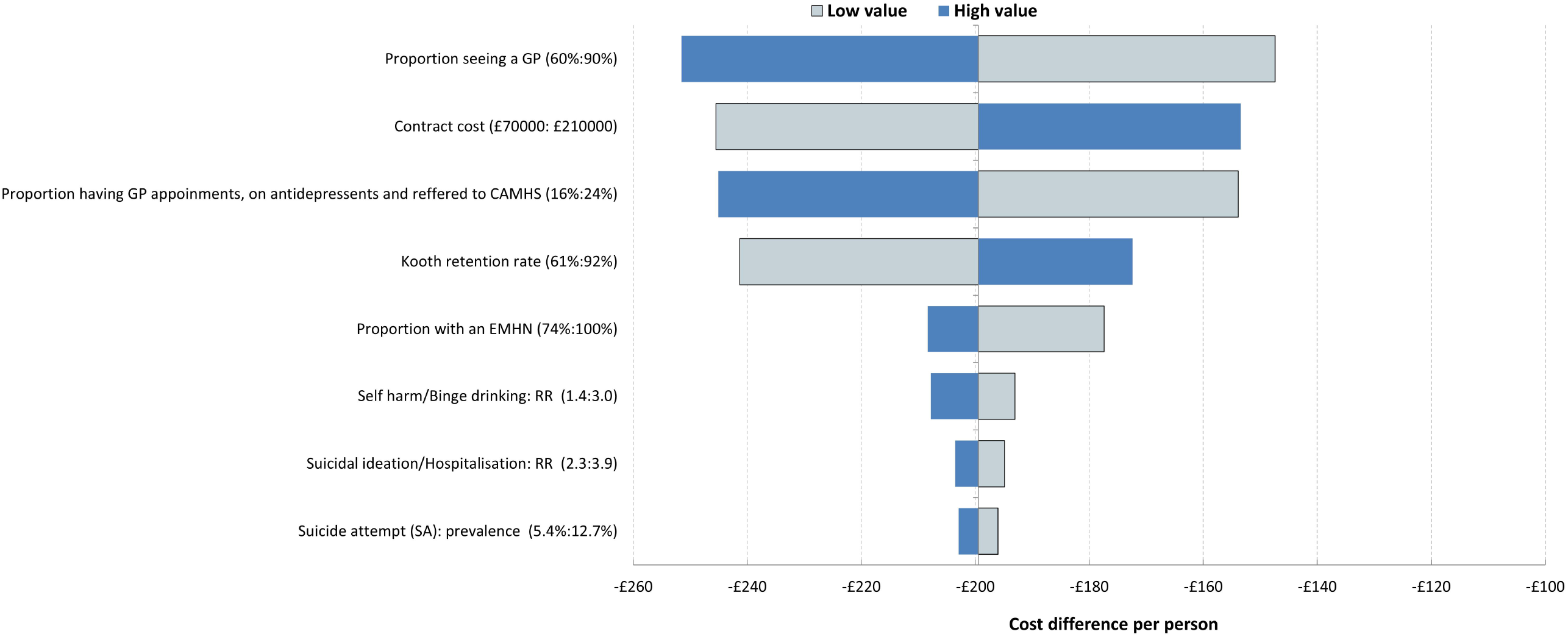

## Discussion

The results suggest that the Kooth mental health platform could be a cost-saving intervention for CYP with EMHN. This early economic model demonstrates a one-year cost saving to the UK government of £199 per engaged user. Engagement with Kooth could result in the reduction of societal events for CYP, such as hospitalisations, committing crimes, smoking and binge drinking. Clinical experts validated the assumption that engaging with a mental health platform, such as Kooth, may also result in a reduction in GP attendances and antidepressant prescriptions. These assumptions were tested using DSA which found Kooth to remain cost saving to UK public services.

Throughout the development of the economic model, the authors identified a lack of published evidence which evaluates the impact of mental health digital platforms on CYP. A systematic review conducted in 2020 identified eleven randomised control trials exploring the impact of mental health apps on CYP (29). All of the included studies looked at the immediate term impact of the app, with the longest follow up period being 6 months (30). The National Institute for Health and Care Excellence (NICE) published guidelines on behaviour change through digital and mobile health interventions (31). In these guidelines NICE emphasised the need for evidence-based behaviour change techniques and the benefits of digital health interventions as a supplement to existing services. Therefore, in sharing these study findings it is hoped that it will contribute to, and encourage, a wider body of published literature in this direction.

In 2013 Clayton and Illback published a paper which aimed to economically justify Jigsaw, a mental health service for CYP in Ireland (32). Like our paper, Clayton and Illback aimed to evaluate the cost impact of early intervention and prevention in CYP. In the analysis they assumed a 5% reduction in psychiatric medication prescriptions in all youth and a 20% reduction in the cost attributable to GP and primary care mental health services, as a result of Jigsaw. These alone totalled 3 million euros in cost savings. Although these figures are based on assumptions, they can be used to support our findings that providing mental health support for CYP could reduce the demand on primary services and reliance on medication.

The impact of poor mental health on educational attainment was explored in the model, however a monetary value was not attached to this outcome given the uncertainty beyond a one-year time horizon. The development of depression and self-harm during primary and secondary education has been previously linked to a decline in future educational attainment of young people (33).

Further, impact of online mental health platforms for young people could help to reduce burden on existing face to face health services. Digital access may overcome some of the barriers in attending in-person GP sessions during working hours and with regards to equitable access of GPs, promoting wider access to mental health support (34).

### Strengths and weaknesses

EMHN was determined through the YP-CORE scores of Kooth users at the time of registration. The YP-CORE score falls into different thresholds, healthy (0–5), low (6–10), mild (11–14), moderate (15– 19), moderate-to-severe (20–24), and severe (25 and above) (7). Scores above the moderate threshold were used as a proxy for EMHN. However, online mental health platforms could also impact those with a low score on the YP-Core, these young people were not captured in the model.

The model focussed on a one-year time horizon. The short time horizon was due to only short term data of one month being published in the LSE study (8). Extending the time horizon of the model beyond one year would introduce further uncertainty in the analysis. However, if changes in immediate outcomes, following engagement with the Kooth platform, were to remain beyond one year the model will underestimate the true cost savings.

Given a lack of evidence linking immediate to intermediate outcomes for CYP in the literature, the economic model focussed on the impact of the Kooth platform on SDQ, self-harm and suicidal ideation. However, it has been shown that the platform can also have a positive impact on other factors, including worries about COVID-19, psychological distress, hope, arguments with parents, closeness to parents and loneliness (8). Therefore, cost savings are likely to be underestimated due to the model focusing on a subset of CYP mental health outcomes.

The model required a large number of assumptions in terms of inputs. These inputs were clinically validated by two clinical practitioners external to the service, who work in the field of CYP’s mental health. These uncertainties were explored in the DSA to determine how varying these inputs would impact on the cost difference. For all parameters, the cost difference remained in the -£140 to -£260 per person range and Kooth remained cost saving.

In the model the impact of Kooth on mental health cannot be separated from the impact of any additional services used by CYP. Young people using Kooth may have access to additional services for their mental health. For example, school counsellors or general practitioners. Furthermore, it was assumed that changes in immediate outcomes are only shown in the EMHN CYP who have already ‘presented’ in this issue while previously using the Kooth platform.

The model primarily evaluated quantitative published evidence and is therefore not able to capture all decision-making discussion points. Patient choice should always be qualitatively considered, this being in line with the emphasis on patient empowerment from the NHS long term plan (35). Evidence was not available to apply utility values and hence quality adjust life years (QALYs) to the immediate mental health outcomes.

## Conclusion

The economic case for online mental health platforms, such as Kooth, as an additional resource alongside traditional mental health services should be given further consideration. This study found a one-year cost saving to the UK government of £199 per engaged user when Kooth is made available to young people. Analysis into the impact of online mental health services on young people is recommended to reduce current uncertainties and assumptions.

## Supporting information

Reporting guidelines

Conflict of interest

## Data Availability

Data that are publicly available have been cited in the reference list, data not publicly available are available upon reasonable request to the authors

## Acknowledgements

We would acknowledge Tom Kayll, Harry Maher and Ellen Howard for support with data extraction from Kooth and for their useful discussions during the study. We would also like to acknowledge Charlotte Mindel, Gareth Evans and Andreas Paris for their useful discussions during the study. Lastly, we would like to acknowledge the clinical team within Kooth Plc led by Dr. Hannah Wilson who provided useful feedback about the model inputs.

