## Supplementary data for "An economic evaluation of Kooth, a web-based mental health platform for children and young people with emerging mental health needs"

Smoking comorbidity calculations

| **Smoking comorbidity** | **RR** | |
| --- | --- | --- |
|  | **Males** | **Females** |
| Stroke(1) | 1.47 | 2.03 |
| Lung cancer(2) | 23.6 | 7.8 |
| Myocardial infarction(3) | 2.23 | 3.46 |
| Coronary heart disease(4) | 1.6 | 1.7 |
| Chronic obstructive pulmonary disease(5) | 7.55 | 3.4 |

RR, Relative risk.

**Baseline costs:**

| **Smoking comorbidity** | **Baseline cost** |
| --- | --- |
| Stroke(6) | £13,902 |
| Lung cancer(7) | £10,031 |
| Myocardial infarction(8) | £4,364 |
| Coronary heart disease(9) | £1,204 |
| Chronic obstructive pulmonary disease(10) | £941 |

Inflated to 2021

**Adjusted cost calculations:**

Baseline prevalence of comorbidities are needed to determine prevalence of smokers with comorbidities.

**Baseline prevalence and sources:**

| **Non-smoking comorbidity prevalence** | **Males** | **Females** |
| --- | --- | --- |
| **Stroke** | | |
| Age 12-15(11) | 0.34% | 0.34% |
| Age 16-24(11) | 0.34% | 0.34% |
| **Lung cancer** | | |
| Age 12-15(12) | 0.01% | 0.01% |
| Age 16-24(12) | 0.01% | 0.01% |
| **Myocardial infarction** | | |
| Age 12-15(13) | 0.00% | 0.00% |
| Age 16-24(13) | 0.00% | 0.26% |
| **Coronary heart disease** | | |
| Age 12-15(14) | 0.00% | 0.00% |
| Age 16-24(14) | 0.31% | 0.31% |
| **Chronic obstructive pulmonary disease** | | |
| Age 12-15(15) | 0.31% | 0.31% |
| Age 16-24(15) | 3.86% | 3.86% |

**Non-smoker baseline prevalence multiplied by RR to estimate smoker prevalence.**

| **Calculated smoking comorbidity prevalence** | **Males** | **Females** |
| --- | --- | --- |
| **Stroke** | | |
| Age 12-15 | 0.5% | 0.69% |
| Age 16-24 | 0.5% | 0.69% |
| **Lung cancer** | | |
| Age 12-15 | 0.15% | 0.07% |
| Age 16-24 | 0.15% | 0.07% |
| **Myocardial infarction** | | |
| Age 12-15 | 0.00% | 0.00% |
| Age 16-24 | 0.00% | 0.90% |
| **Coronary heart disease** | | |
| Age 12-15 | 0.00% | 0.00% |
| Age 16-24 | 0.49% | 0.53% |
| **Chronic obstructive pulmonary disease** | | |
| Age 12-15 | 2.34% | 1.05% |
| Age 16-24 | 29.17% | 13.40% |

The 2019/20 ONS statistics on the UK population show a 30.11% and 69.89% split for 12-15 years and 16-24 years for UK males. The 2019/20 ONS statistics on the UK population show a 30.24% and 69.76% split for 12-15 years and 16-24 years for UK females.

The 2019/20 ONS statistics on the UK population show a 51.37% and 48.63% split between males (5,203,477) and females (4,926,394) between 12-24 years.

These weightings were used to calculate total age (12-24 years) and gender (male and female) percentage in each smoking comorbidity.

**Adjusted percentage smokers:**

| **Smoking comorbidity** | **Adjusted percentage smokers:** |
| --- | --- |
| Stroke | 0.25% |
| Lung cancer | 0.10% |
| Myocardial infarction | 12.65% |
| Coronary heart disease | 0.22% |
| Chronic obstructive pulmonary disease | 0.14% |

Adjusted percentage changes between non-smokers and smokers were multiplied by annual comorbidity costs to calculate the adjusted annual comorbidity cost difference between smokers and non-smokers used in the economic model.

| **Smoking comorbidity** | **RR** | | **Cost** | |
| --- | --- | --- | --- | --- |
|  | **Males** | **Females** | **Baseline** | **Adjusted** |
| Stroke | 1.47 | 2.03 | £13,902 | £82.38 |
| Lung cancer | 23.6 | 7.8 | £10,031 | £11.08 |
| Myocardial infarction | 2.23 | 3.46 | £4,364 | £13.32 |
| Coronary heart disease | 1.6 | 1.7 | £1,204 | £4.29 |
| Chronic obstructive pulmonary disease | 7.55 | 3.4 | £941 | £145.33 |
| **Total** | | | | £256.40 |

RR, Relative risk.

Binge drinking calculations

**The problematic drinking population, monthly prevalence:**

| **Percentage of the problematic drinking population:** | **Monthly** |
| --- | --- |
| Committing a crime(16) | 4.3% |
| Hospitalised(16) | 1.00% |
| Having unprotected sex(17) | 1.10% |

**The problematic drinking population, associated cost:**

| **Cost per event** | **Cost** |
| --- | --- |
| Committing a crime(17) | £1,143 |
| Hospitalised(18) | £849 |
| Having unprotected sex(18) | £68 |

Average yearly cost of a problematic drinking individual were calculated by multiplying monthly prevalence by cost and scaling up to one year.

| **Calculated average yearly cost of problematic drinking individual** | **Monthly cost** |
| --- | --- |
| Committing a crime | £49.15 |
| Hospitalised | £8.49 |
| Having unprotected sex | £0.75 |
| Total monthly | £58.39 |
| **Total yearly** | **£700.64** |
